# Short Report: Investigating Oropouche as a possible etiology for fevers of unknown origin in a clinical cohort from Colombia, 2013-2015

**DOI:** 10.1101/2024.11.06.24316756

**Authors:** Christine S. Walsh, Jenny C. Cardenas, Lady Y. Gutierréz-Silva, Maria U. González, Christopher N. Mores, Berlin Londono-Renteria, Rebecca C. Christofferson

## Abstract

Currently, the largest outbreak of Oropouche virus (OROV) in Latin America and the Caribbean, has also spread across the globe to Italy, Spain, and Germany, bringing this virus into the public domain. With over 8,000 cases and counting in Brazil, there have been two deaths in young women with no known comorbidities, and infection with Oropouche was associated with a late term stillbirth. While OROV has been sporadically detected in Colombia since 2017, with a handful of cases in Cúcuta and Leticia, the distribution of OROV prior to that remains unknown. Therefore, we undertook to test a clinical cohort of febrile patients from Colombia to determine if OROV was circulating within the human population earlier than previously determined. We screened 631 serum samples collected in 2014-2015 from three municipalities: Cúcuta, Los Patio, and Ocaña. We found evidence of OROV infection in three patients from Ocaña, though live virus was not recoverable from patient sera. These data suggest that OROV circulation, while sporadically detected in Colombia, has circulated earlier than previously thought. With the emergence and expansion of new or neglected viruses, there is a need to expand screening of febrile patients both retro- and prospectively to describe and better understand the distribution of arbovirus circulation in human populations. Surveillance for OROV across South America is necessary going forward, especially as the outbreak in Brazil continues.

## Introduction

Arboviruses are a cause of public health concern due to the variety of clinical symptoms ranging from mild febrile illness to death [1]. Zika virus (ZIKV), dengue virus (DENV), Chikungunya (CHIKV) virus, and yellow fever virus (YFV) are well-known arboviruses that can result in a number of human illnesses [1]. These viruses can have similar symptomology spectrum, which includes a mild febrile illness [2, 3]. A lack of resources available to diagnose and treat patients can also lead to misdiagnosis or an ultimate lack of definitive diagnosis. In these cases, diagnosis may be a non-descript diagnosis of fever of unknown origin [4].

Currently, the largest outbreak of OROV in Latin America and the Caribbean, primarily Cuba and Brazil [5] has also been detected in returning travelers across the globe, such as in Italy, Spain, the United States, and Germany [6-9]. With over 10,000 cases as of the writing of this article, there have been two deaths in young women with no known comorbidities [10, 11]. Further, infection with OROV has been associated with pregnancy loss [11, 12] which led officials to warn doctors that this could be a contributor to poor pregnancy outcomes, including microcephaly [13, 14].

OROV has been found in human patients in numerous countries that share border with Colombia: Ecuador, Peru, Brazil [15, 16]. Although there have been only a handful of documented cases of OROV in Colombia since 2017 [17-19], the presence of *Culicoides paraensis*, the primary vector of OROV in Colombia suggests opportunity for robust transmission [20]. In Colombia, the detected distribution of OROV has been limited, with cases in Turbaco (n=1) in 2017, in Cúcuta (n=1) and Leticia (n=3) in 2019-22 [17-19].We undertook to test an earlier clinical cohort of febrile patients from Colombia collected between 2013-2015 to determine if OROV was circulating within the human population earlier than previously determined.

## Materials and Methods

### Patient samples

Serum samples were obtained between 2013 and 2015 from patients in Colombia at E.S.E. Hospital Universitario Erasmo Meoz, E.S.E. Hospital de Los Patios, E.S.E. Hospital Emiro Quintero Cañizares, from the municipalities of Cúcuta, Los Patios, and Ocaña, respectively (LSU IRB protocol #3415). All patients were tested for OROV. Available demographics include location and year of collection, sex and age.

### RNA Extraction and qRT-PCR

RNA was extracted using the KingFisher (ThermoFisher) robot and the Ambion MagMag viral isolation kit (ThermoFisher), per manufacturer’s instructions. Extracted RNA samples were then tested for the presence of OROV RNA using the OROV S segment previously published [21]. The sequences are as follows: forward primer 5’ TCCGGAGGCAGCATATGTG 3’, reverse primer 5’ ACAACACCAGCATTGAGCACTT 3’, probe 5’ (FAM) CATTTGAAGCTAGATACGG 3’. Samples were run on the Roche LightCycler 96. Samples with a Cq value of ≤ 40 were considered positive and curves were visualized to ensure no false signal. OROV cycling parameters for the S segment were reverse transcription at 50°C for 300 seconds, then 95°C for 20 seconds, and 2-step amplification at 45 cycles of 95°C for 3 seconds and 60°C for 30 seconds [21].

## Results

### Evidence of acute OROV infection in 2015 in a clinical cohort

Human serum samples were collected from three cities in the Department of Norte de Santander, Colombia (**Figure 1**), which borders Venezuela to the east. Cúcuta is an urban area in a valley of the Colombian Andes sitting at 320 meters above sea level (masl), with a population of over 770,000 and is situated approximately 250 masl [22]. Los Patios is a suburban area outside of Cúcuta with a population of just over 77,000 people [23], and is part of the large metropolitan area of Cúcuta. Ocaña is a more elevated municipality in the west of the Department at an elevation of 1200 masl and is more rural than the other two sites [24].

**Figure 1:**
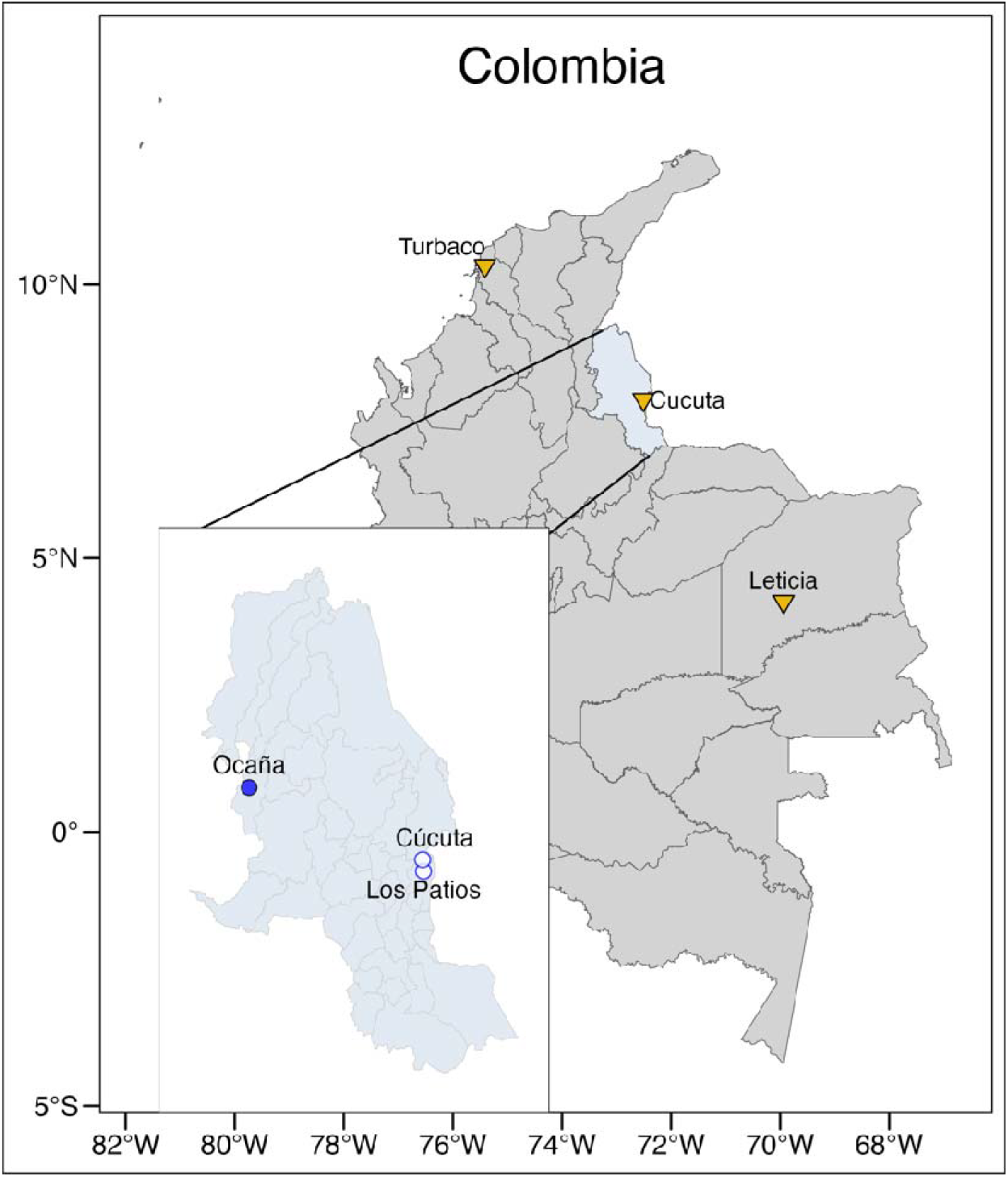
Location of previously detected OROV cases (yellow triangles) prior to 2017. Location of the municipalities from which patients were sampled in Norte de Santander, Colombia (circles). Ocaña (filled blue circle) had 3 cases of OROV in 2015.

A total of 631 samples were collected, 177 from Cúcuta, 189 from Los Patios, and 265 from Ocaña. The sex distribution was approximately equal with 304 males (48.3%) and 316 females (50.1%); 11 individuals had no data regarding sex. Of the 631 samples, 246 samples (39.0%) were collected in 2014 period, while 383 (60.7%) were collected in 2015. Two samples were missing collection dates. Detailed sample demographics are described in **Table 1**. The age distribution is given in Table 1, but 166 (26.3%) were between 0-15 years of age, 180 (28.5%) were between 16-30, 70 (11.1%) were between 31-45, 86 (13.6%) were between 46-60, and 118 (18.7%) were 61 years or older. The median age was 12 years old, with a range of 0.11-92 years of age (IQR: 7 – 24.5 years).

**Table 1.** Human Serum Sample Demographics.

|  | Cúcuta<br>(n=177) | Los Patios<br>(n=189) | Ocaña<br>(n=265) | Total<br>(n=631) |
| --- | --- | --- | --- | --- |
| **Sex** |  |  |  |  |
| Female | 87 | 86 | 145 | 316 |
| Male | 89 | 102 | 113 | 304 |
| Unknown | 1 | 3 | 7 | 11 |
| **Age** |  |  |  |  |
| Unknown | 1 | 4 | 7 | 12 |
| 0-5 | 50 | 35 | 26 | 111 |
| 6-15 | 103 | 63 | 77 | 243 |
| 16-25 | 14 | 35 | 67 | 116 |
| 26-35 | 4 | 12 | 28 | 44 |
| 36-45 | 2 | 12 | 30 | 44 |
| 46-55 | 2 | 4 | 12 | 18 |
| 56-65 | 0 | 12 | 7 | 18 |
| 66+ | 1 | 12 | 77 | 24 |
| **Collection Year** |  |  |  |  |
| 2014 | 0 | 132 | 114 | 246 |
| 2015 | 177 | 57 | 149 | 383 |
| Unknown | 0 | 0 | 2 | 2 |

Three individuals tested positive for OROV via qRT-PCR. All samples were collected in Ocaña in 2015 (**Figure 1**). One patient was a child < 5 years old and another in the 6-15 age range, both males. The third was a female in the 31-45 age range. While qRT-PCR detected OROV, attempts to grow the virus in Vero cells was unsuccessful, indicating that the infection was perhaps in late, clearance phase. This is the first indication that OROV circulates this west in Colombia.

## Discussion

The clinical manifestation during the early stages of OROV infection mimic those of other etiologies endemic in Latin America [3]. Many pathogens cause febrile illness in the Colombian population, including yellow fever, Zika, DENV, chikungunya, and Mayaro, and Venezuelan equine encephalitis viruses. All of these are arboviruses transmitted by arthropod vectors and can result in febrile illness [2]. Given the relatively common set of symptoms and a lack of robust surveillance and diagnostic frameworks targeted OROV, there is ample opportunity for misdiagnosis and underreporting [15]. Because of the non-specific generalized febrile illness associated with the majority of OROV infections, it is important to understand the epidemiology of this previously non-descript infection to determine whether historical outbreaks have gone undetected. Our data indicate that OROV has been circulating in Colombia earlier than previously detected and is the furthest west that OROV has been detected within Colombia. This is important for understanding the force of introductions – past and future – to anticipate future outbreaks.

The region of Norte se Santander is characterized for a high volume of exchange of people and animals between Colombia and Venezuela [25, 26]. This area has perceived a significant increase in cases of vector-borne diseases such as malaria and dengue in the past decade. Interestingly, Norte de Santander was the first department where Zika was first detected in the Country between 2015 and 2015 and reported some of the higher rates of infections during the outbreak in the country [27]. In the case of DENV, this area is among the Colombian departments with the highest number of dengue fever cases annually [28, 29] with four serotypes circulating at a given time [30]. A recent phylogeographical analysis suggested a significant diffusion route of DENV between Colombia and Venezuela [31]. Hence, with this study we aimed to evaluate the potential for OROV to circulate in the area at earlies points of time. By increasing surveillance for neglected arboviruses in human, vector, and animal populations, we can close some of the knowledge gaps regarding neglected tropical disease epidemiology including transmission, distribution, and characteristic disease symptoms. This could also inform future preventative and preparedness measures for future outbreaks of emerging viruses.

Our data suggest that OROV circulation has been sporadic and limited in Colombia, though that it has been present in the human population since at least 2015. As the crow flies, Ocaña is 325 km southeast from Turbaco, 1437 km northwest of Leticia, and 101 northwest of Cucùta. This represents the most westward case of OROV in the country and could indicate that a wider risk exists in the country that may not be linked to trade or border-adjacent transmission.

Careful surveillance is needed going forward, especially as the outbreak in Brazil continues. With the emergence and expansion of new or neglected viruses, there is a need to expand screening of febrile patients both retro- and prospectively to describe and better understand the distribution of arbovirus circulation in human populations. This would lead to a more comprehensive view of disease burden resulting from a specific viruses, which would then inform efforts and resource allocation for surveillance and prevention programs [32].

## Data Availability

All data produced in the present work are contained in the manuscript

## Acknowledgements

The authors thank the population of Norte de Santander for supporting this study. We also thank the Hospitals involved reviewing and approving the protocols of this study and allowing access to the clinical laboratories. Further, we’d like to thank Snowflake and Tina for their role as placeholders and critics. And to Dr. Roland Cooper.

